# Uncertainty Quantification in Epidemiological Models for COVID-19 Pandemic

**DOI:** 10.1101/2020.05.30.20117754

**Authors:** Leila Taghizadeh, Ahmad Karimi, Clemens Heitzinger

**Affiliations:** Institute for Analysis and Scientific Computing, Vienna University of Technology, Vienna, Austria; School of Mathematical and Statistical Sciences, Arizona State University, Tempe, AZ 85287

**Keywords:** Coronavirus forecasting, epidemic models, COVID-19 pandemic, Bayesian inversion, MCMC methods, data science

## Abstract

The main goal of this paper is to develop the forward and inverse modeling of the Coronavirus (COVID-19) pandemic using novel computational methodologies in order to accurately estimate and predict the pandemic. This leads to governmental decisions support in implementing effective protective measures and prevention of new outbreaks. To this end, we use the logistic equation and the SIR system of ordinary differential equations to model the spread of the COVID-19 pandemic. For the inverse modeling, we propose Bayesian inversion techniques, which are robust and reliable approaches, in order to estimate the unknown parameters of the epidemiological models. We use an adaptive Markov-chain Monte-Carlo (MCMC) algorithm for the estimation of a posteriori probability distribution and confidence intervals for the unknown model parameters as well as for the reproduction number. Furthermore, we present a fatality analysis for COVID-19 in Austria, which is also of importance for governmental protective decision making. We perform our analyses on the publicly available data for Austria to estimate the main epidemiological model parameters and to study the effectiveness of the protective measures by the Austrian government. The estimated parameters and the analysis of fatalities provide useful information for decision makers and makes it possible to perform more realistic forecasts of future outbreaks.

## 1 Introduction

The Coronavirus COVID-19 pandemic is a new infectious disease which emerged from China in fall 2019 and then spread around the world. This pandemic spreads through (micro-) droplets and its outbreak speed is very high.

The first reported case of SARS-CoV-2 was identified in Wuhan, China. The first case outside of China was reported in Thailand on 13 January 2020 [1]. Since then, this ongoing outbreak has now spread all over the world [2]. So far (at the time of writing), this pandemic has infected around 5 230 000 individuals around the world and caused more than 335 000 deaths. Out of more than 2 780 000 active cases around the world, 2% are critical patients. Source of the data is the Johns Hopkins CSSE database (https://github.com/CSSEGISandData/COVID-19).

The COVID-19 pandemic was confirmed to have spread to Austria on 25 February 2020 by a 24-year-old man and a 24-year-old woman (according to the Federal Ministry of Social Affairs, Health, Care and Consumer Protection, Republic of Austria, https://info.gesundheitsministerium.at) traveling from Lombardy, Italy, who were treated at a hospital in Innsbruck. So far in Austria more than 16 400 people have been infected and 633 deaths have been reported. Furthermore, out of 833 active cases, 4% are critical patients. Figure 1 displays daily confirmed cases in Austria and Figure 2 illustrates total cumulative count of confirmed and active cases in Austria. By removing deaths and recoveries from total cases, we obtain the “currently infected cases” or “active cases” (cases still awaiting an outcome).

**Figure 1:**
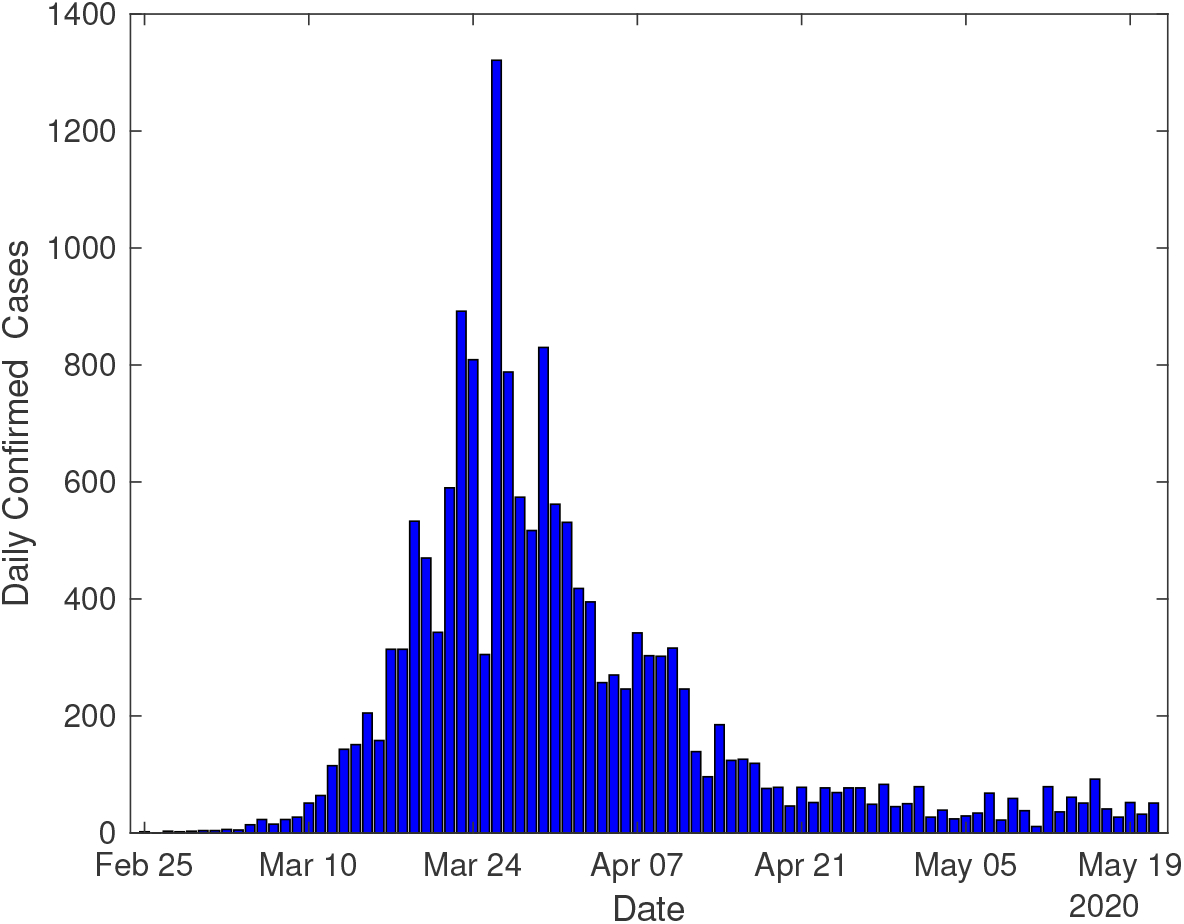
Daily confirmed count of coronavirus infected cases (till May 21st) in Austria.

**Figure 2:**
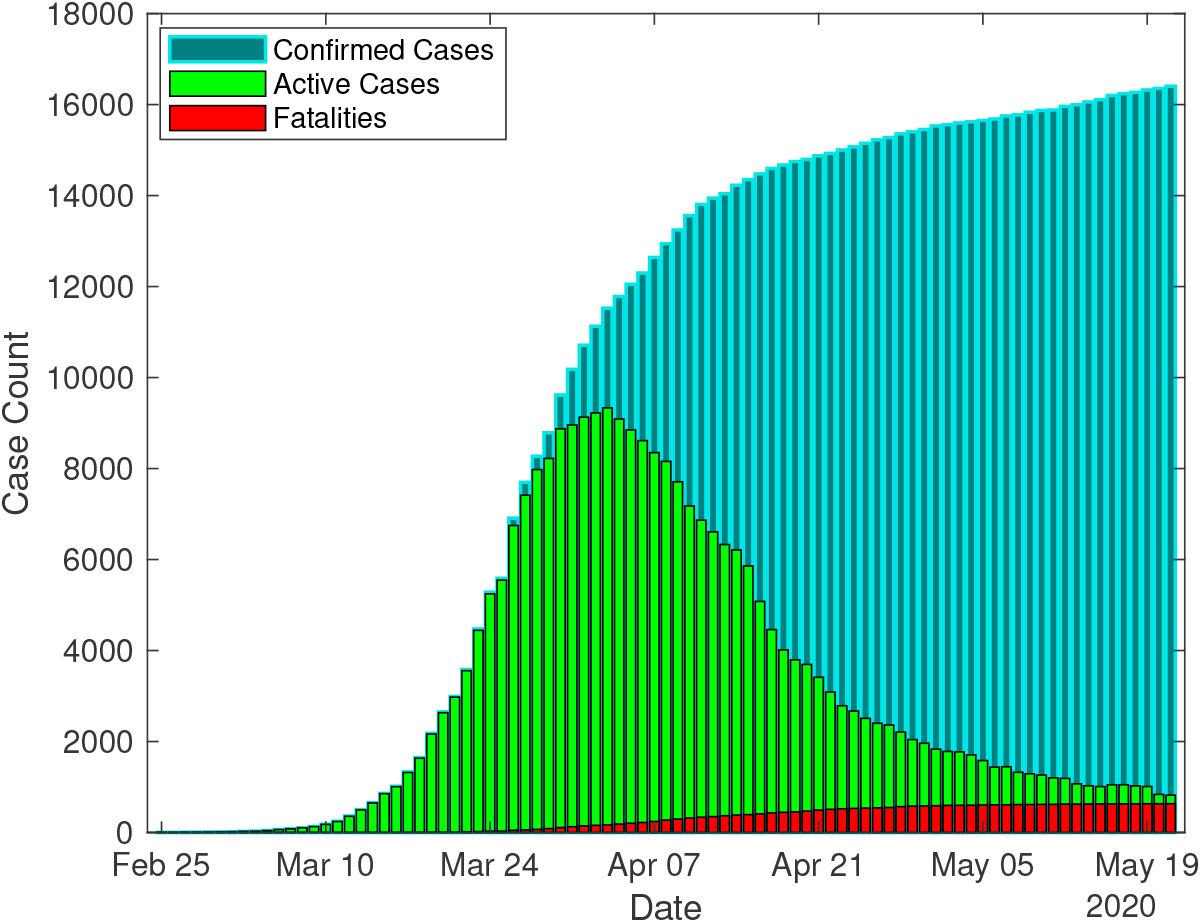
Total cumulative count of Coronavirus confirmed infected case and active cases (till May 21st) in Austria.

Infected people need breathing assistance and a large number of them require medical treatment in an intensive care unit (ICU). Countries which are affected by COVID-19 attempt to keep the daily number of cases below the capacity of their health care system. In order to avert the disastrous inundation of hospitals, the virus must be kept from spreading fast. To this end, countries have been implementing protective measures such as closing schools, canceling mass gatherings, working from home (home office), self-quarantine, self-isolation, avoiding crowds, social distancing, wearing protection masks, etc.

In this work, we propose Bayesian inference for the analysis of the COVID19 data in order to estimate the crucial unknown quantities of the pandemic models. We use an adaptive MCMC method to find the probability distributions and confidence intervals of the epidemiological models parameters using the Austrian infection data. We use this analysis for the prediction of the duration of the epidemic in Austria as well as the total number of infected people and fatalities till the end of the epidemic. The model validation shows a very good agreement between the computational and measurement data of infections in Austria which proves the reliability and the accurateness of the predictions. This is of great importance for making governmental decisions in implementing the measures in order to prevent the spread of the virus.

This paper is organized as follows: Section 2 presents the logistic and

SIR epidemiological models and introduces their unknown parameters. Section 3 is devoted to the Bayesian analysis as the inversion method which is proposed for quantifying the uncertain model parameters in the epidemiological models. Numerical results of the forward and inverse epidemic models including the quantifying the models uncertain parameters, models validation using the measurement data, pandemic forecast, fatality analysis and the effect of governmental protective measures are presented in Section 4. Finally, conclusions are drawn in Section 5.

## 2 Mathematical Models for COVID-19

Predictive mathematical models are essential for the quantitative understanding of epidemics and for supporting decision makers in order to implement the most effective and protective measures. Many mathematical models for the spread of infectious diseases [3–6] and in particular for the novel COVID-19 [7–11] have been presented and analyzed. Here we start with the logistic equation as a preliminary model for epidemics and continue with the SIR model and its extensions [12–17].

### 2.1 The Logistic Model

The logistic equation is a nonlinear ordinary differential equation, which is used for modeling population growth. This ODE is also well-known as logistic growth model and is given by

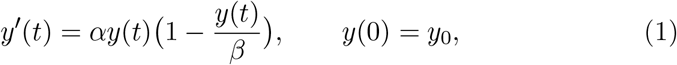

where *y*_0_ ≠ 0 is the initial population size (initial number of confirmed cases), *y* denotes population size (total accumulated confirmed cases) and *t* time. Furthermore, *α* and *β* are respectively the growth rate (infection rate) and the carrying capacity (maximum number of confirmed cases), which are positive constants.

The solution to the logistic model equation is

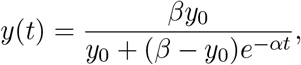

which can be rewritten as where

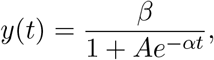

where

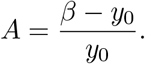

Furthermore, the expected peak date of the outbreak is calculated as

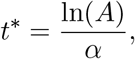

which is the time when the expected maximal rate of confirmed cases (growth rate) occurs.

### 2.2 The SIR model

The susceptible-infected-recovered (SIR) model is an epidemiological model that computes the number of people infected with a contagious disease in a closed population over time. The Kermack-McKendrick model is one of the SIR models, which is defined by the system

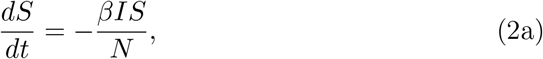

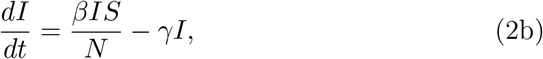

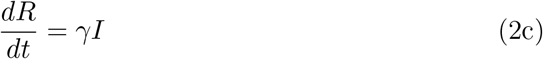

of ordinary differential equations, where *β* and *γ* are the infection and recovery rates, respectively. The model consists of three components: *S* for the number of susceptible, *I* for the number of infectious, and *R* for the number of recovered or deceased (or immune) individuals. Furthermore, *N* denotes the constancy of population, i.e.

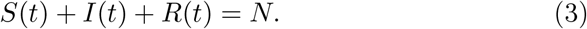

Moreover, the dynamics of the infectious class depends on the reproduction number/ratio, which is defined as

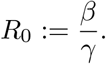

If the reproduction number is high, the probability of a pandemic is high, too. This number is also used to estimate the herd immune threshold (HIT). If the reproduction number multiplied by the percentage of susceptible individuals is equal to 1, it shows an equilibrium state and thus the number of infectious people is constant.

Additionally, the recovery period is defined by

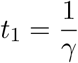

and describes the average days to recover from infection. The transmission period in the sense of the average days to transmit the infection to a person is defined by

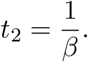

However in a population with vital dynamics, new births can provide more susceptible individuals to the population, which sustain an epidemic or allow new introductions to spread in the population. Taking the vital dynamics into account, the SIR model is extended to

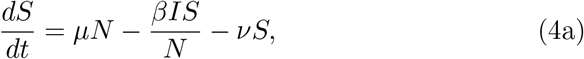

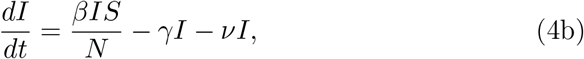

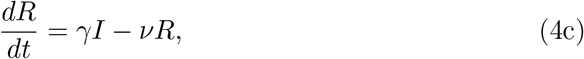

where *μ* and *ν* denote the birth and death rates, respectively. To maintain a constant population, we assume *μ* = *ν* is the natural mortality rate.

## 3 Bayesian Inversion for the Model Parameters

We propose Bayesian inversion methods in order to solve the backward/inverse problem of COVID-19, which is the problem of accurate estimation of the epidemiological model parameters as well as the reproduction ratio. Bayesian inference in the context of the statistical inversion theory is based on Bayes’ Theorem and, compared to traditional inverse methods, has the advantage of updating the prior knowledge about the unknown quantity using the measurement/observation data. Furthermore, Bayesian analysis is a robust inversion technique for parameter extraction and gives the (a posteriori) probability distribution and confidence intervals for the unknowns instead of providing a single estimate. We have already successfully applied Bayesian inversion techniques to various PDE models in engineering and medicine in order to identify parameters (see for instance [18–21]).

### 3.1 Bayesian Analysis

As mentioned, the Bayesian inversion approach is a robust and reliable technique to quantify the uncertain parameters of the epidemic models. In fact, the solution of the inverse problem is the posterior density that best reflects the distribution of the parameter based on the observations. As the observations or measurements are subject to noise, and the observational noise, i.e., the error *e* due to modeling and measurement, is unbiased and iid, it can be represented by random variables as

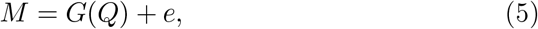

where *e* is a mean-zero random variable and *M* is a given random variable representing observed data or measurements, for which we have a model *G*(*Q*) (observation operator) dependent on a random variable *Q* with realizations *q* = *Q*(*ω*) representing parameters to be estimated [22].

Assume a given probability space (Ω*, F, P*), where Ω is the set of elementary events (sample space), *F* a *σ*-algebra of events, and *P* a probability measure. Furthermore assume that all the random variables are absolutely continuous.

Bayes’ Theorem in terms of probability densities can be written as with

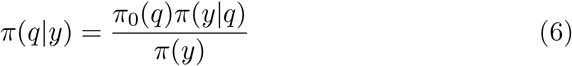

with

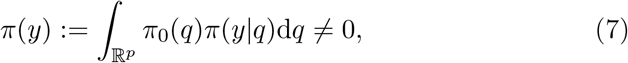

where the unknown parameters *q* = (*q*_1_*,…, q_p_*) ∈ ℛ^*p*^ and the observed data *y* are realizations of the random variables *Q* and *M*, respectively. Furthermore, *π*_0_(*q*), *π*(*q*∣*y*), and *π*(*y*∣*q*) are the probability density functions of the prior, posterior, and (data) sampling distributions, respectively. The density *π*(*y*∣*q*) of the data provides information from the measurement data to update the prior knowledge, and it is well-known as the likelihood density function. The goal of Bayesian inversion is to estimate the posterior probability density function *π*(*q*∣*y*), which reflects the uncertainty about the quantity of interest *q* using measurement data *y*.

Equation (1) gives the posterior density and summarizes our beliefs about *q* after we have observed *y*. Therefore, Bayes’ Theorem for inverse problems can be stated as follows.

**Theorem 1** (Bayes’ Theorem for inverse problems [22, 23]). *Let π*_0_(*q*) *be the prior probability density function of the realizations q of the random parameter Q. Let y be a realization or measurement of the random observation variable M. Then the posterior density of Q given the measurements y is*

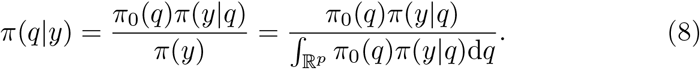

Computing the integral appearing in Bayes’ Theorem 1 is costly especially if the parameter space ℛ*^p^* is high-dimensional. Another problem with quadrature rules is that they require a relatively good knowledge of the support of the probability distribution, which is usually part of the information that we seek [22, 23]. In Section 3.2 we shortly discuss the algorithms for Bayesian estimation, which do not require evaluations of the integral and that are used to achieve the numerical results for the nonlinear model equation.

### 3.2 Markov-Chain Monte-Carlo (MCMC) Methods

Markov-chain Monte-Carlo methods are a class of Monte-Carlo methods with the general idea of constructing Markov chains whose stationary distribution is the posterior density [22]. The Metropolis-Hastings algorithm is an MCMC algorithm to draw samples from a desired distribution. In this algorithm, the first state of the chain *q*_0_ is chosen and then the new state *q_k_, k* = 1, 2,…, *N*, of the chain is constructed based on the previous state *q_k_*_−1_. To this end, a new value *q*^∗^ using the proposal density function *J* (*q*^∗^∣*q_k_*_−1_) is proposed. Admissibility of this proposed value is tested by means of calculating the acceptance ratio *α*(*q*^∗^ *q_k_*_−1_). If the proposed value is admissible, it is accepted as *q_k_*, otherwise the old value is kept and a new proposal is made. For more details about MCMC methods see for example [24–27].

Although the convergence speed is determined by the choice of a good proposal distribution, at least tens or hundreds of thousands of samples are necessary to converge to the target distribution. Choosing the optimal proposal scaling is a crucial issue and affects the MCMC results; if the covariance of the proposal distribution is too small, the generated Markov chain moves too slowly, and if it is too large, the proposals are rejected. Hence, optimal proposal values should be found to avoid both extremes, which leads to adaptive MCMC methods [28–30]. In the following section, we will consider an adaptive algorithm that helps sample from potentially complicated distributions.

### 3.3 Delayed-Rejection Adaptive-Metropolis (DRAM) algorithm

Searching for a good proposal value can be done manually through trial and error, but this becomes intractable in high dimensions. Therefore, adaptive algorithms that find optimal proposal scales automatically are advantageous. The delayed-rejection adaptive-Metropolis (DRAM) algorithm is an efficient adaptive MCMC algorithm [29]. It is based on the combination of two powerful ideas to modify the Markov-chain Monte-Carlo method, namely adaptive Metroplolis (AM) [31,HYPERLINK \l “_bookmark45” 32] and delayed-rejection (DR) [33,HYPERLINK \l “_bookmark47” 34], which are used as global and local adaptive algorithms, respectively. AM finds an optimal proposal scale and updates the proposal covariance matrix, while DR updates the proposal value when *q*^∗^ is rejected.

The basic idea of the DR algorithm is that, if the proposal *q*^∗^ is rejected, delayed rejection (DR) provides an alternative candidate *q*^∗∗^ as a secondstage move rather than just retaining the previous value *q_k_*_−1_. This process is called delayed rejection, which can be done for one or many stages. Furthermore, the acceptance probability of the new candidate(s) is calculated. Therefore, in the DR process, the previous state of the chain is updated using the optimal parameter scale or proposal covariance matrix that has been calculated via the AM algorithm.

The AM algorithm is a global adaptive strategy, where a recursive relation is used to update the proposal covariance matrix. In this algorithm, we take the Gaussian proposal centered at the current state of the chain *q_k_* and update the chain covariance matrix at the *k*-th step using

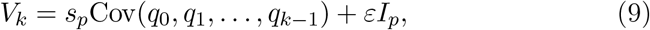

where *s_p_* is a design parameter and depends only on the dimension *p* of the parameter space. This parameter is specified as *s_p_*: = 2.38^2^/*p* as the common choice for Gaussian targets and proposals [35], as it optimizes the mixing properties of the Metropolis-Hastings search in the case of Gaussians. Furthermore, *I_p_* denotes the *p*-dimensional identity matrix, and *ε >* 0 is a very small constant to ensure that *V_k_* is not singular, and in most cases it can be set to zero [29].

The adaptive Metropolis algorithm employs the recursive relation

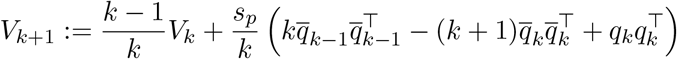

to update the proposal covariance matrix, where the sample mean 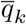 is calculated recursively by

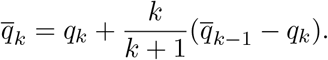

A second-stage candidate *q*^∗∗^ is chosen using the proposal function

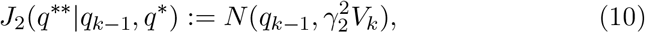

where *V_k_* is the covariance matrix produced by the adaptive algorithm (AM) as the covariance of the first-stage and *γ*_2_ < 1 is a constant. The probability of accepting the second-stage candidate, having started at *q_k_*_−1_ and rejected *q*^∗^, is

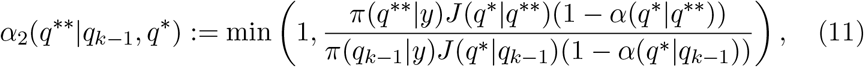

where *α* is the acceptance probability ((12)) in the non-adaptive approach. The acceptance probability is computed so that reversibility of the posterior Markov chain is preserved (for more details see for example [22, §8.6]). The DRAM technique is summarized in Algorithm 1.

## 4 Numerical Results

In this section, we present simulation results of Bayesian inversion and the adaptive MCMC method (see Algorithm 1) for the two epidemic models, namely the logistic and the SIR models, using the data of the COVID-19 outbreak in Austria. The results include model parameter estimation, model validation and outbreak forecasting.

### Algorithm 1 The DRAM algorithm

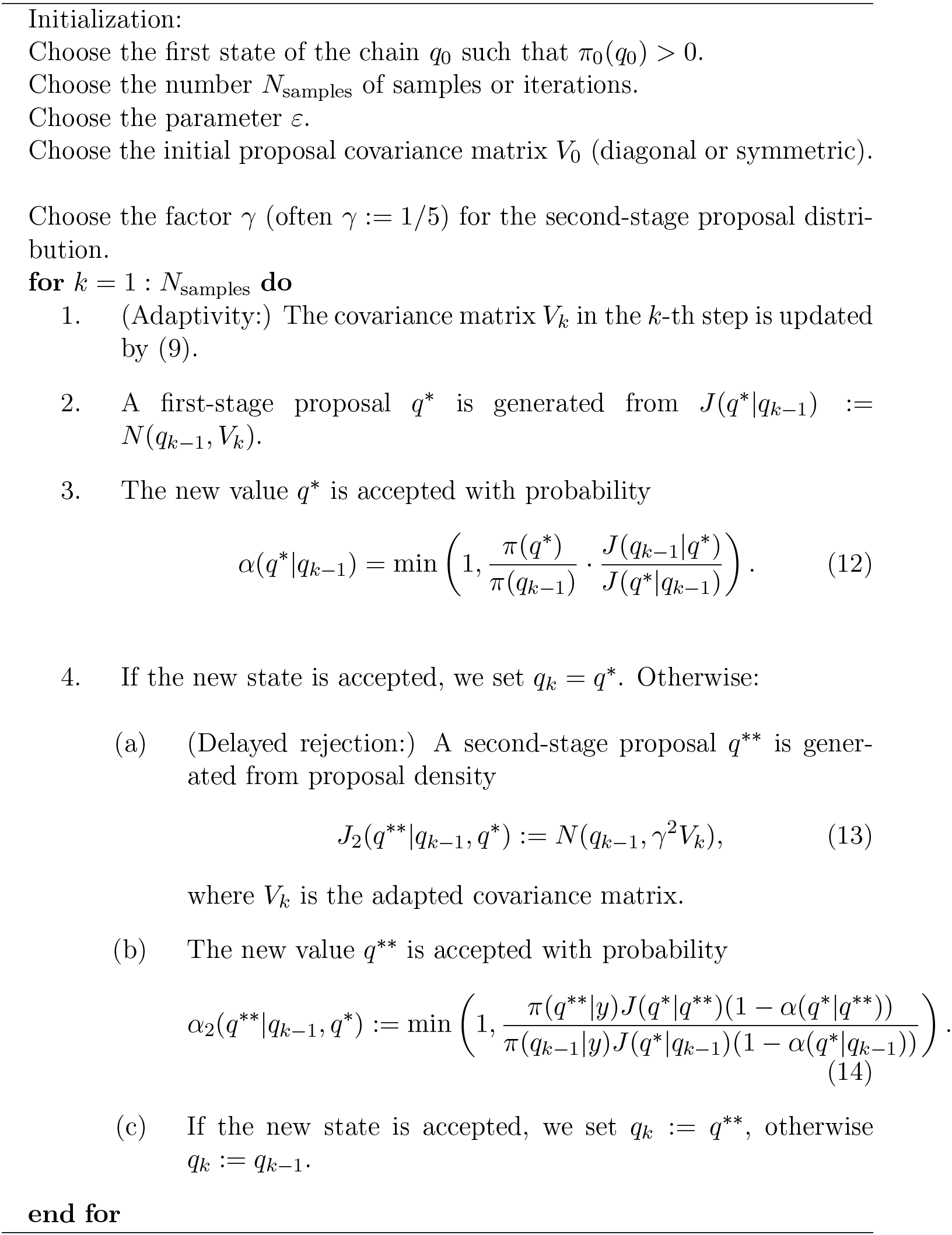

### 4.1 Parameter Estimation

According to Bayesian analysis, the unknown parameters of the logistic and SIR models using the data of COVID-19 outbreak in Austria were found and summarized in Table 1 and Table 3, respectively. These tables show the confidence intervals for the models parameters as well as the mean of the obtained Markov chains in the Bayesian inference.

**Table 1:** Estimated confidence intervals and mean of Markov chains for the parameters of the logistic model using Bayesian inversion method for Austria.

| Parameter | Description | Estimated mean | Confidence interval 95% |
| --- | --- | --- | --- |
| $\alpha$ | Growth rate | 0.28 | [0.23, 0.33] |
| $\beta$ | Carrying capacity | 14 974 | [12 703, 17 244] |

**Table 2:** Estimated peak time for Austria using the logistic model.

| Quantity | Description | Estimation |
| --- | --- | --- |
| $t^*$ | Peak time | March 26-27 |

**Table 3:** Estimated confidence intervals and means of Markov chains for the parameters of the SIR model using Bayesian inversion method for Austria.

| Parameter | Description | Estimated mean | Confidence interval 95% |
| --- | --- | --- | --- |
| $\beta$ | Transmission rate | 0.36 | [0.32, 0.39] |
| $\gamma$ | Recovery rate | 0.06 | [0.03, 0.09] |
| $N$ | Total population | 17 617 | [12 217, 23 016] |

Furthermore, Table 2 and Table 4 include temporal quantities such as peak time of the outbreak estimated using the Bayesian inference for the logistic and SIR models.

**Table 4:**
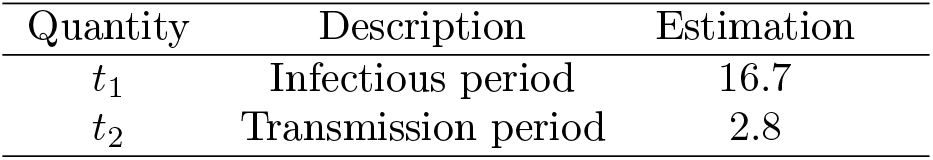
Estimated temporal quantities for Austria using the SIR model.

| Quantity | Description | Estimation |
| --- | --- | --- |
| $t_1$ | Infectious period | 16.7 |
| $t_2$ | Transmission period | 2.8 |

Figure 4 illustrates marginal histograms of posterior distribution for the four quantities of interest in the SIR model, namely *β, γ, N* and *R*_0_ using Bayesian analysis.

**Figure 3:**
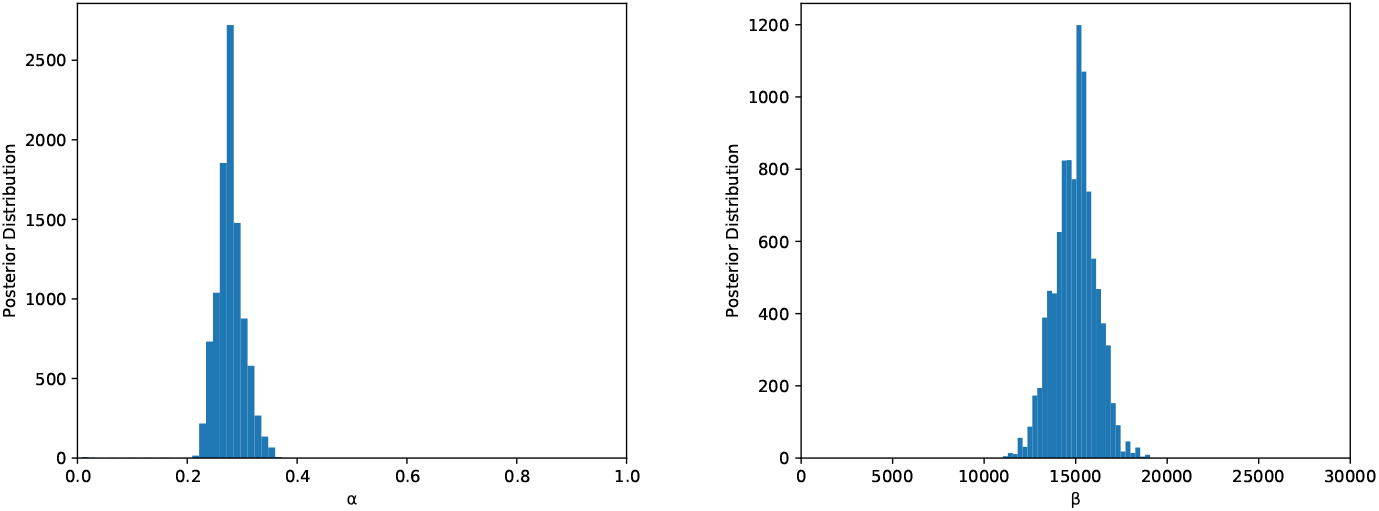
The marginal histograms of posterior distribution for the two quantities of interest in the logistic model, namely *α* and *β*.

**Figure 4:**
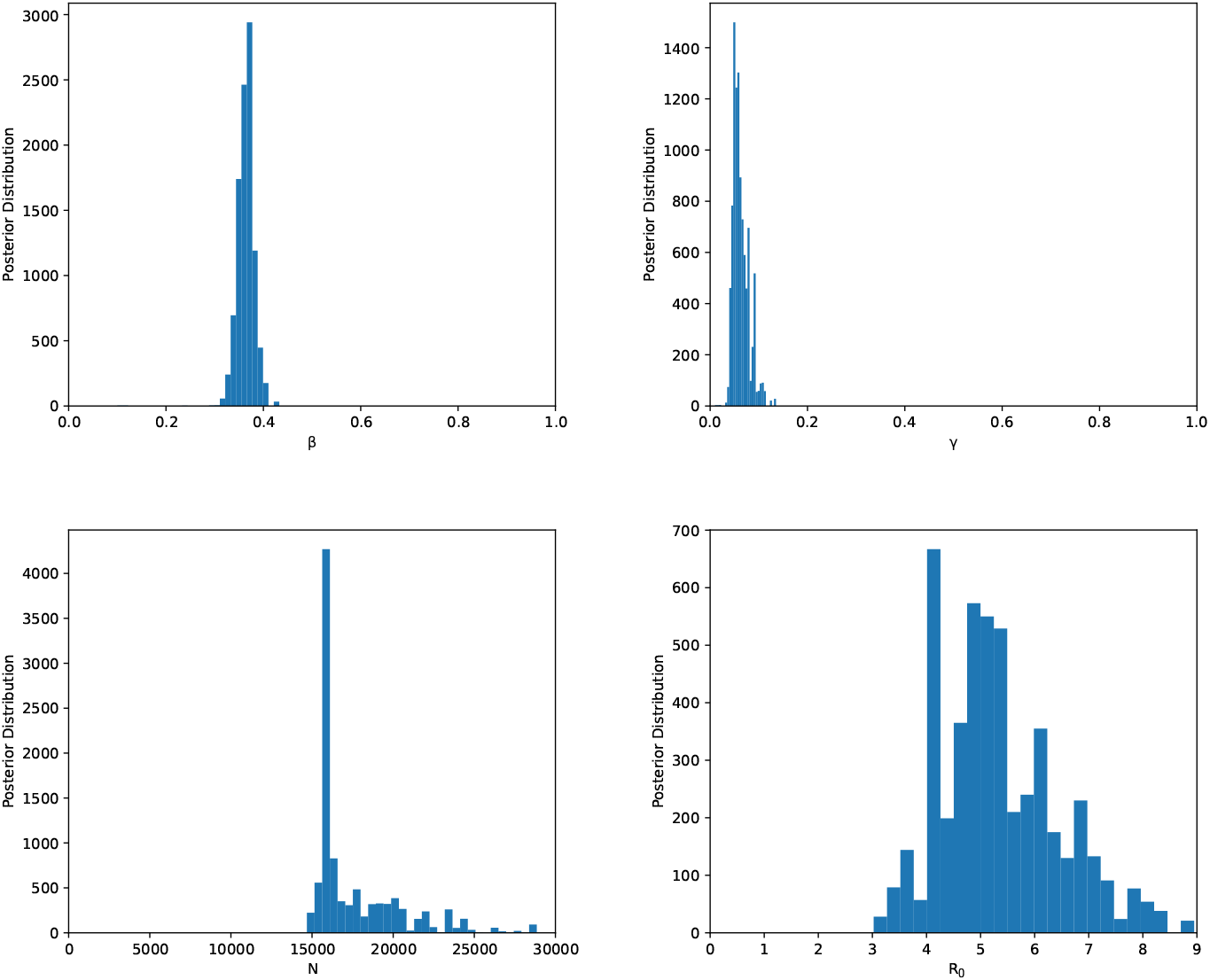
The marginal histograms of posterior distribution for the four quantities of interest in the SIR model, namely *β, γ, N* and *R*_0_.

### 4.2 Model Validation

Here, we aim to evaluate the logistic and SIR models for forecasting the COVID-19 outbreak by comparing the simulation results and the reported data. Figure 5 illustrates the number of infected individuals in Austria till now, as well as our prediction of the number of infected people for the coming days. This prediction is according to the Bayesian inversion for the logistic equation as the epidemic model.

**Figure 5:**
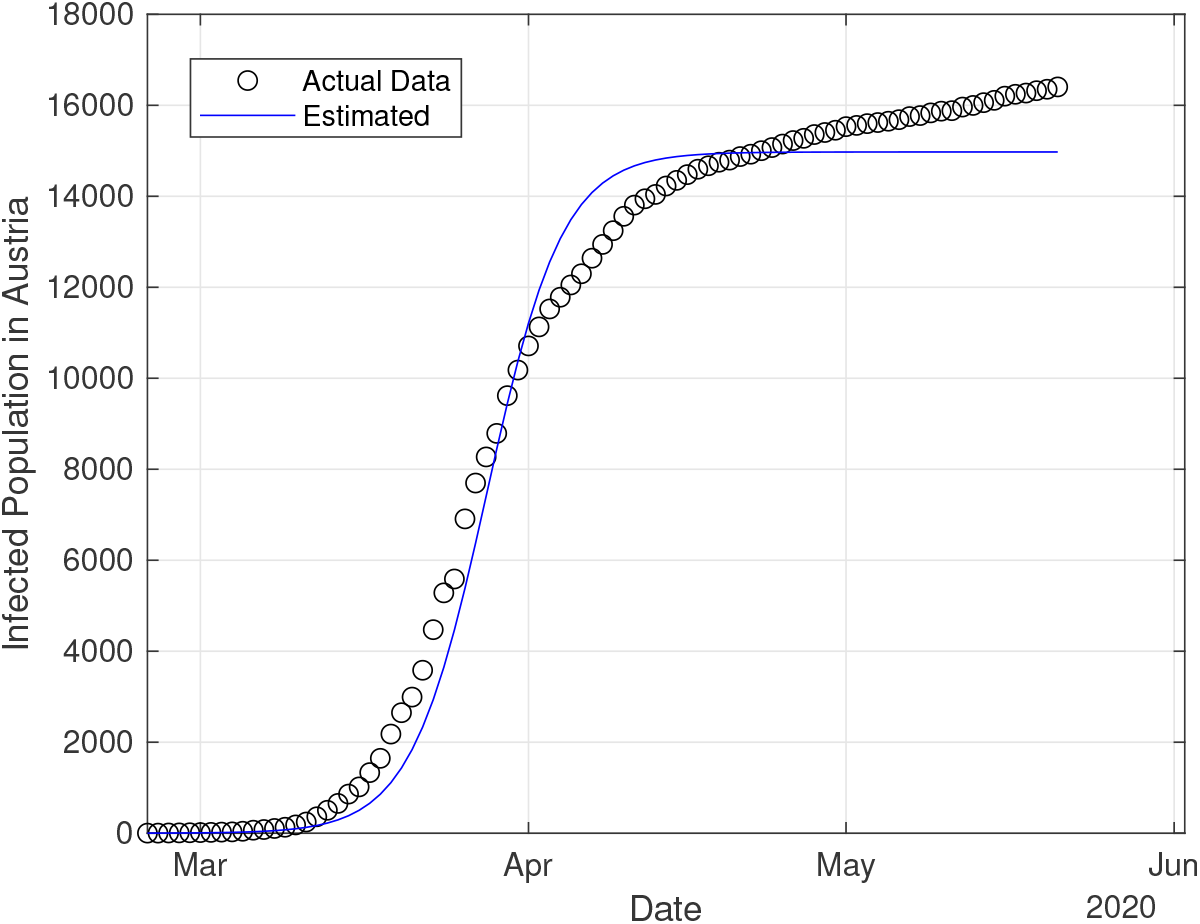
Estimated total cumulative count of coronavirus confirmed infected cases in Austria using Bayesian analysis for the logistic model versus actual or measured infected population.

Now we use the SIR model and estimate the unknown quantities of this epidemic model using Bayesian inference for the spread of the Coronavirus. Quantifying the uncertain parameters such as the reproduction number leads to calculate the average number of days to recover from the infection and gives useful information about properly and accurately implementing protective measures in order to prevent the spread of the virus. Furthermore, the parameter identification in the epidemic model makes it possible to predict the length of the pandemic, the number of infected individuals and the fatality rate.

Figure 6 displays a similar prediction using the Bayesian inference for the SIR model, which shows a very good agreement between the measurements and the simulation. In Figure 7, the actual and estimated infection rates are depicted. This rate is defined by

**Figure 6:**
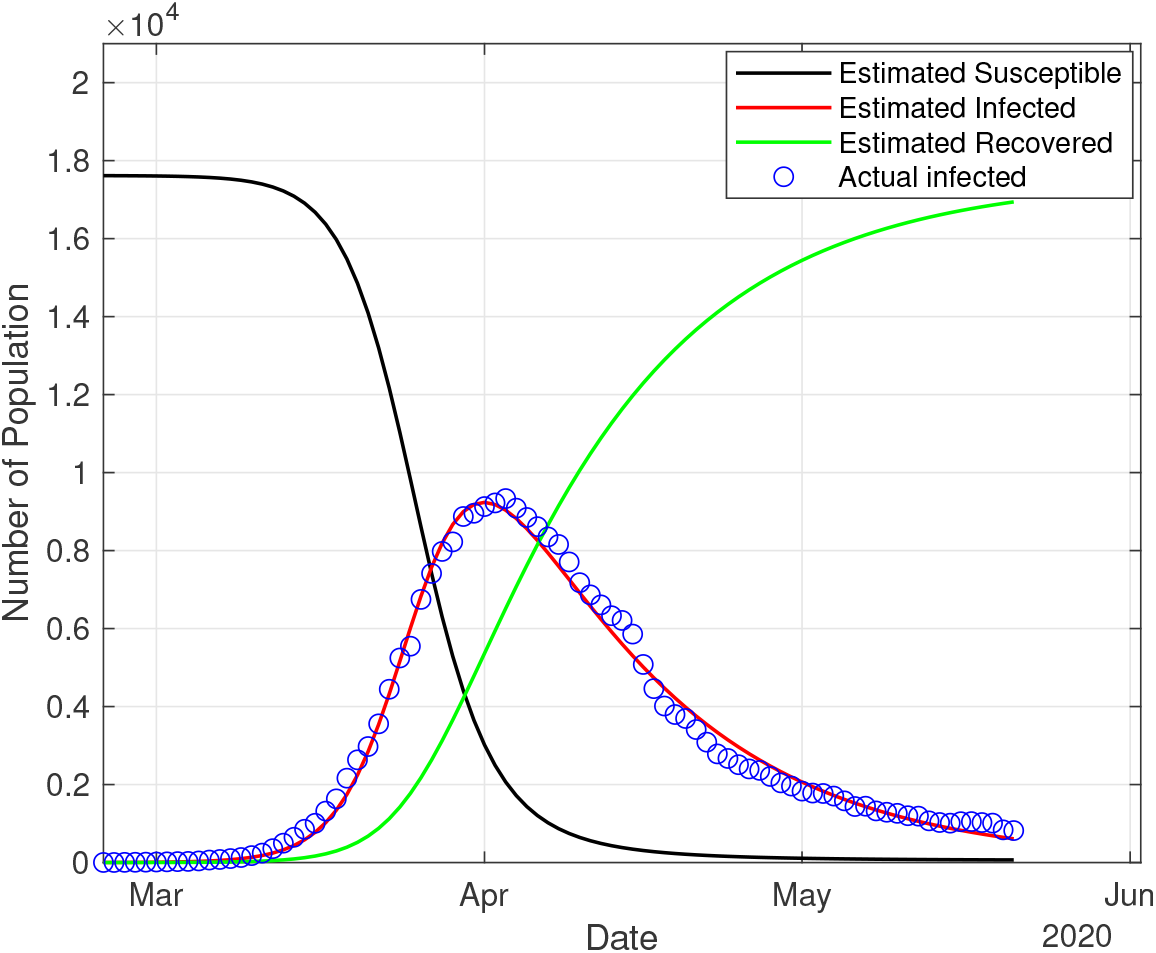
Estimated total count of coronavirus susceptible, infected and recovered cases using Bayesian analysis for the SIR model as well as actual confirmed active cases in Austria.

**Figure 7:**
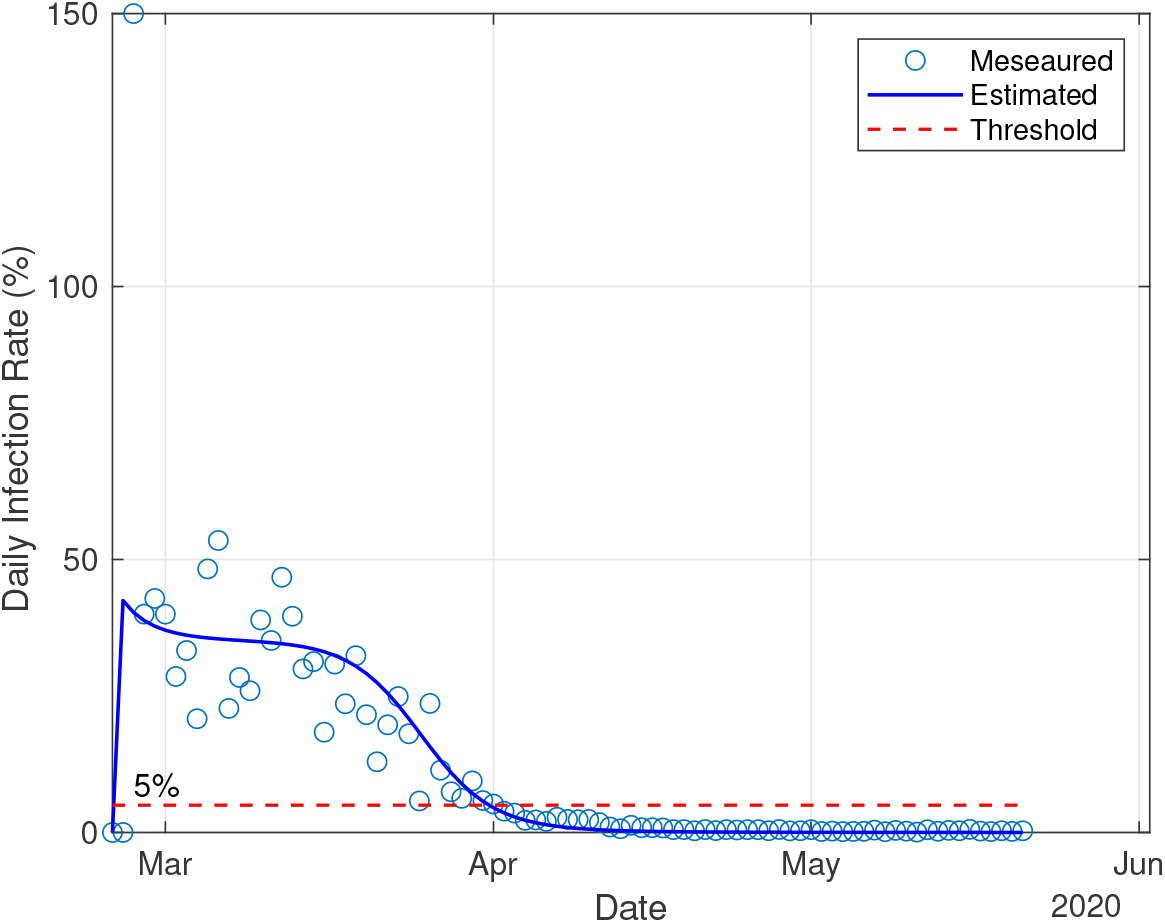
Infection rate in Austria.

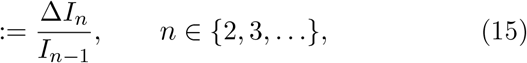

where ∆*I_n_* = *I_n_ I_n_*_−1_, *I_n_* and *I_n_*_−1_ are infected population of consequent times (e.g. in days or weeks), which are obtained using the estimated infection from the SIR model. Figure 8 shows the estimated and actual reproduction number for Austria during the infection time. The first recovery in Austria was reported on March 26, where the reproduction number is estimated as 3. This quantity decays to 1 on April 1, and afterwards remains below 1. In this figure, the threshold of *R* = 1 is also displayed in the sense that there is no immediate public health emergency any more when the reproduction number is below this threshold.

**Figure 8:**
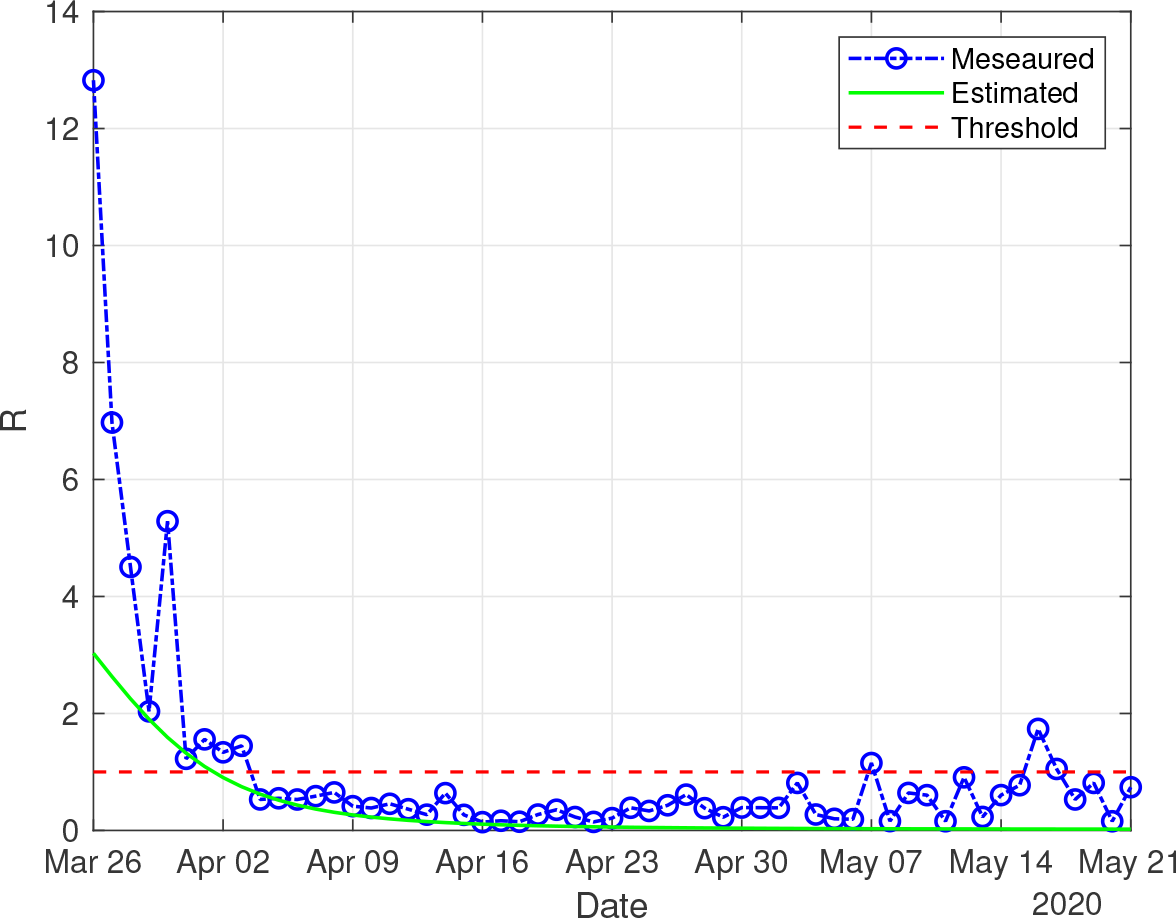
Reproduction number in Austria.

### 4.3 Fatality Analysis

Social-distancing measures started in Austria around March 16, 2020 and improvements were first observed around March 21, which is consistent with the incubation time. In Figure 9, the daily fatalities in Austria as well as the relative change in fatalities are illustrated.

**Figure 9:**
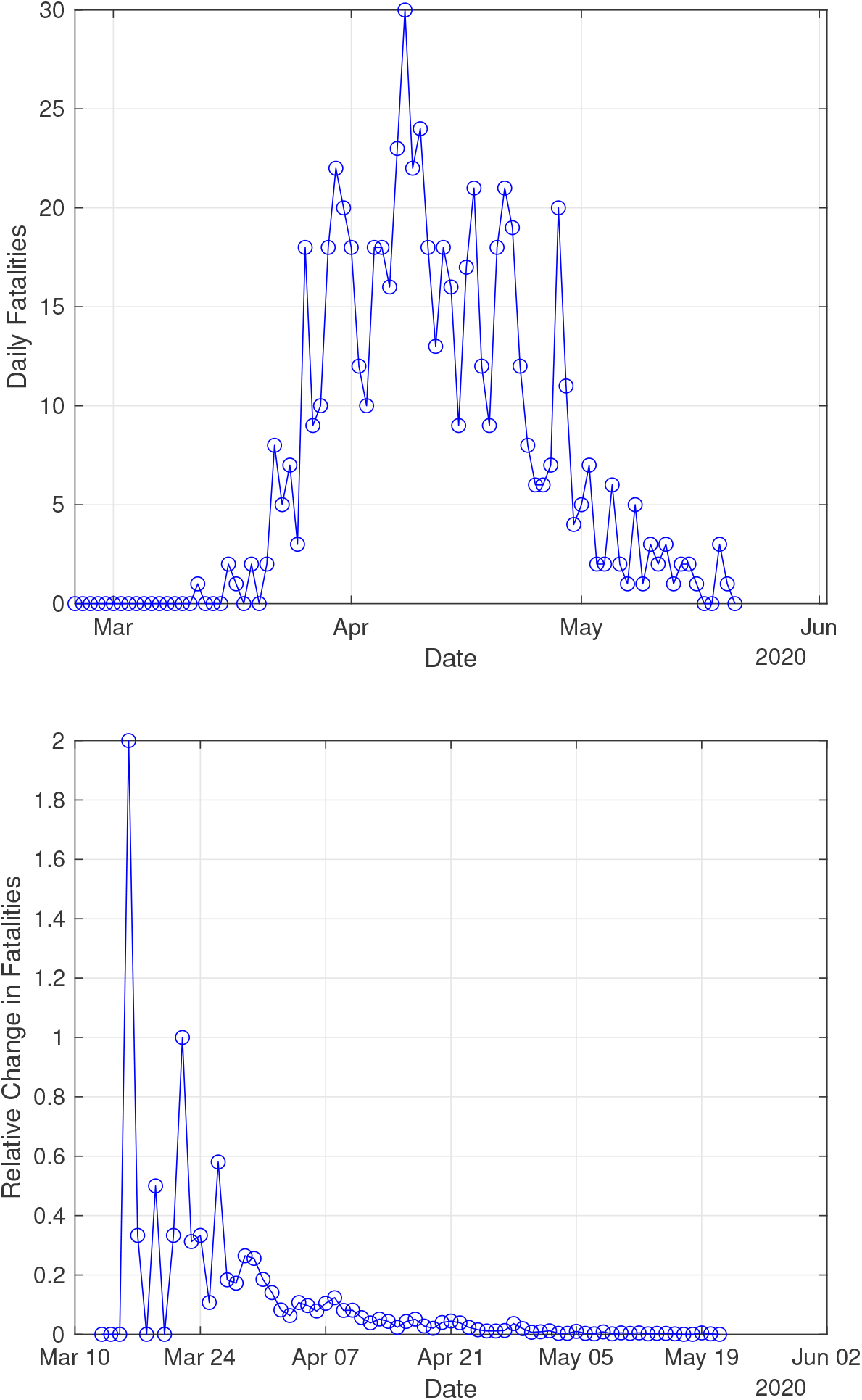
Fatalities in Austria including the daily fatalities (top) and the relative change in fatalities (bottom).

Applying the fatality ratio as well as confirmed infected cases, we present a fatality analysis which is of importance for governmental protective decision making. In epidemiology, a case fatality rate (CFR) is the proportion of deaths from a certain disease compared to the total number of people diagnosed with the disease for a certain period of time. Figure 10 depicts the case fatality rate (CFR), which is defined by

**Figure 10:**
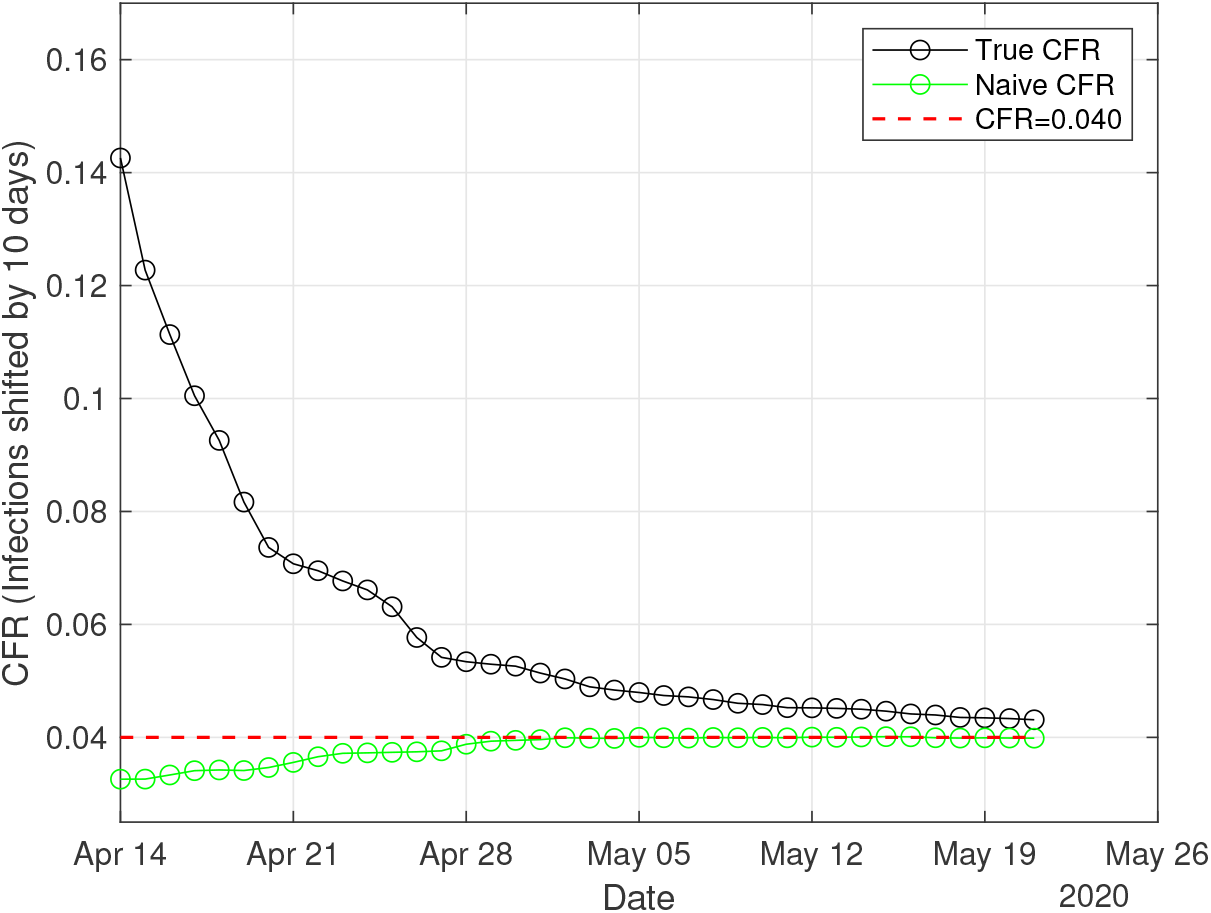
Case fatality rate (CFR) in Austria.

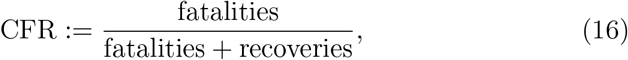

and we call it the true CFR here, in contrast to the naive CFR defined by

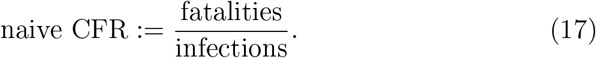

The straightforward calculations using the recorded data in Austria show that both CFR and naive CFR converge to the same value of CFR^∗^ = 0.04 (Figure 10).

We can roughly predict the fatalities using the confirmed infections, a shift (the number of days between confirmation of infection and death), and the CFR. The shift is approximately equal to the time between infection and death (currently estimated to be 18 days) minus the incubation time (currently estimated to be up to 7 days) minus time for testing and reporting (see Figure 11). The estimator of fatalities in Austria is defined by

**Figure 11:**
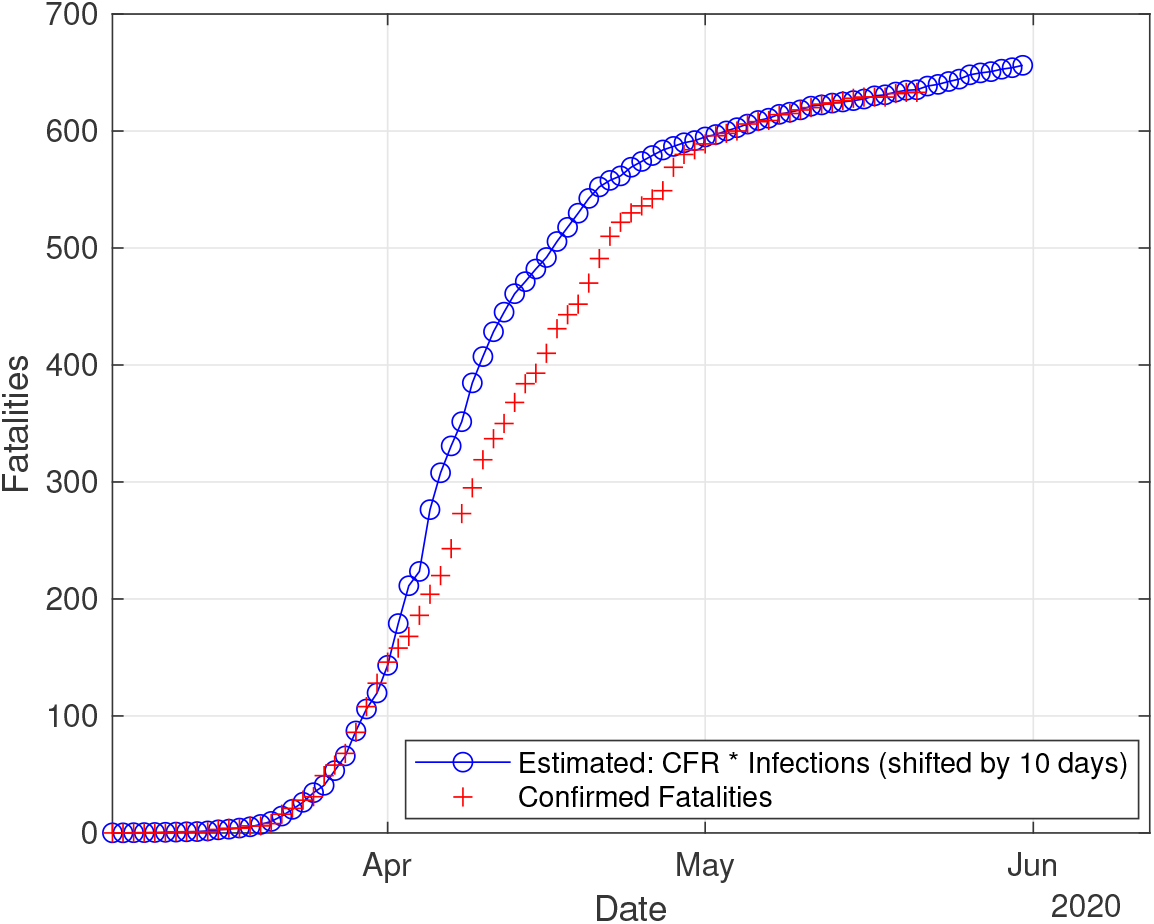
Prediction of number of fatalities in Austria.

fatality cases: = CFR^∗^ × confirmed infected cases (shifted by 10 days).

Figure 11 shows a good agreement between the estimated fatalities and true values in Austria.

### 4.4 The Impact of Governmental Protective Measures

Here, in order to study the effect of the protective measures implemented by the Austrian government, we compare the infection rate and the infected population in different time intervals with and without implementing the measures. Austrian government decided to implement some protective measures such as social distancing around March 10, 2020. Although public health measures were in place from March 16 to control the spread of COVID19, Austrians started to practice social distancing and to wear masks much before. For instance, Austrian universities stopped their physical activities starting March 11 and practices a distance-learning and home-office strategy.

Table 5 shows the weekly infection rate in Austria and how it decays in subsequent weeks. The comparison between the estimated infection rates in subsequent weeks before and after implementing the protective measures highlights the importance and effectiveness of the measures such as socialdistancing and lock-down in controlling and slowing the spread of COVID-19.

**Table 5:**
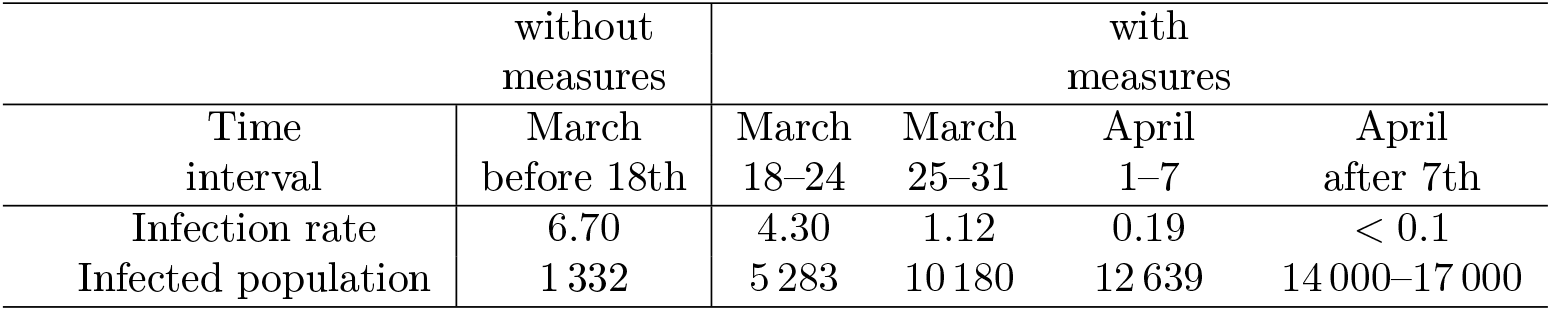
Infection rate (see Equation (15)) and total cumulative infected population at the end of different time intervals with and without implementing the protective measures in Austria.

Furthermore, the fatality forecast in Section 4.3 is valid as long as protective measures are in place, otherwise the number of fatalities will increase due to a large number of infected people and the limit in the capacity of intensive care unit (ICU) beds in Austria as the number of intensive cases increases dramatically. In fact, the main goal of protective measures and lock-down is to “flatten the curve”, i.e., to decrease the infection rate so that the healthcare system is kept from becoming overwhelmed with too many critical cases at the same time. In countries where the counterfactual scenario i.e., no public health interventions is applied, for instance in Sweden, the ICU demand is estimated to be almost 20 times higher than the intensive care capacity in the country and a much larger number of deaths is predicted [36]. If Austrian governmental protective measures would not have taken place, after March 18 the infection rate would have remained almost constant (around 6) and therefore we would expect thousands of infected people and fatalities in Austria.

## 5 Conclusions

In this work, we developed an adaptive Bayesian inversion for epidemiological models, namely the logistic and the SIR models, in order to solve the inverse problem of estimating unknown quantities for the novel Coronavirus COVID19. Quantifying the uncertainties in these models is essential since it leads to describe the characteristics of the epidemics on one hand and accurately forecasting the pandemic on the other hand. The proposed inversion recipe is robust and yields probability distributions and confidence intervals for the unknown parameters of the epidemic models including the growth rate of the outbreak and transmission and recovery rates as well as the reproduction number, whose quantification is crucial.

We applied our methodology to the publicly available data for Austria to estimate the main epidemiological model parameters and to study the effectiveness of the protective measures by the Austrian government as well as fatality analysis. We also validated the presented models by comparing the simulated and measured data, whose results show a very good agreement.

Based on Bayesian analysis for the logistic model, the means of the growth rate *α* and the carrying capacity *β* are estimated respectively as 0.28 and 14 974. Furthermore for these parameters, 95% confidence intervals of [0.23, 0.33] and [12 703, 17 244] are obtained. Moreover for the parameters of the SIR model, namely the transmission rate *β*, recovery rate *γ* and total population *N*, the means of 0.36, 0.06 and 17 617 as well as the 95% confidence intervals of [0.32, 0.39], [0.03, 0.09] and [12 217, 23 016] are inferred. Additionally, we obtained the infectious period of 17 days and transmission period of 3 days for COVID-19 in Austria. The first recovery in Austria was reported on March 26, where the reproduction number is estimated as 3. This quantity decays to 1 on April 1, and since then remains below 1.

Analyzing data of infected, recovered and death cases, we obtained that the case fatality rate (CFR) has converged to the value 4%. This estimation makes it possible to forecast the fatalities in the coming 10 days. According to our analysis, the total number of death in Austria is estimated as 633 in May 21, which perfectly matches the measured data.

Furthermore, we estimated the infection rate for consequent weeks starting from before implementing the protective measures, which shows a significant decay after the measures are in place. If the governmental protective measures would not have been implemented, the estimated infection rate before March 18 would have remained almost constant and thus a thousands of people would have been infected and would have died. These results indicate the impact of the measures such as social distancing and lock-down in controlling the spread of COVID-19.

## Data Availability

the Johns Hopkins CSSE database (https://github.com/CSSEGISandData/COVID-19)

## Acknowledgments

The authors acknowledge support by FWF (Austrian Science Fund) START project no. Y660 *PDE Models for Nanotechnology*.

## Notes

### Competing Interest Statement

The authors have declared no competing interest.

### Author Declarations

There is no ethical consumption.

## References

[1] David S Hui, Esam I Azhar, Tariq A Madani, Francine Ntoumi, Richard Kock, Osman Dar, Giuseppe Ippolito, Timothy D Mchugh, Ziad A Memish, Christian Drosten, et al. The continuing 2019-nCoV epidemic threat of novel coronaviruses to global health—the latest 2019 novel coronavirus outbreak in Wuhan, China. International Journal of Infectious Diseases, 91:264, 2020.

[2] World Health Organization (WHO). Coronavirus Disease 2019 (COVID-19) Situation Report – 97. WHO, 2020.

[3] Roy M Anderson, B Anderson, and Robert M May. Infectious diseases of humans: dynamics and control. Oxford University Press, 1992.

[4] Odo Diekmann and Johan Andre Peter Heesterbeek. Mathematical epidemiology of infectious diseases: model building, analysis and interpretation, volume 5. John Wiley & Sons, 2000.

[5] Herbert W Hethcote. The mathematics of infectious diseases. SIAM Review, 42(4):599–653, 2000.

[6] Fred Brauer, Carlos Castillo-Chavez, and Carlos Castillo-Chavez. Mathematical models in population biology and epidemiology, volume 2. Springer, 2012.

[7] Tao Zhou, Quanhui Liu, Zimo Yang, Jingyi Liao, Kexin Yang, Wei Bai, Xin Lu, and Wei Zhang. Preliminary prediction of the basic reproduction number of the Wuhan novel coronavirus 2019-nCoV. Journal of Evidence-Based Medicine, 2020.

[8] Liangrong Peng, Wuyue Yang, Dongyan Zhang, Changjing Zhuge, and Liu Hong. Epidemic analysis of COVID-19 in China by dynamical modeling. arXiv preprint arXiv:2002.06563, 2020.

[9] Yu Chen, Jin Cheng, Yu Jiang, and Keji Liu. A time delay dynamical model for outbreak of 2019-nCoV and the parameter identification. Journal of Inverse and Ill-posed Problems, 28(2):243–250, 2020.

[10] Igor Nesteruk. Statistics based predictions of coronavirus 2019-nCoV spreading in mainland China. MedRxiv, 2020.

[11] Vivek Verma, Ramesh K Vishwakarma, Anita Verma, Dilip C Nath, and Hafiz TA Khan. Time-to-death approach in revealing chronicity and severity of COVID-19 across the world. PLoS ONE, 15(5):e0233074, 2020.

[12] Gerardo Chowell, Nick W Hengartner, Carlos Castillo-Chavez, Paul W Fenimore, and Jim Michael Hyman. The basic reproductive number of Ebola and the effects of public health measures: the cases of Congo and Uganda. Journal of Theoretical Biology, 229(1):119–126, 2004.

[13] Huwen Wang, Zezhou Wang, Yinqiao Dong, Ruijie Chang, Chen Xu, Xiaoyue Yu, Shuxian Zhang, Lhakpa Tsamlag, Meili Shang, Jinyan Huang, et al. Phase-adjusted estimation of the number of coronavirus disease 2019 cases in Wuhan, China. Cell Discovery, 6(1):1–8, 2020.

[14] W Jumpen, B Wiwatanapataphee, YH Wu, and IM Tang. A SEIQR model for pandemic influenza and its parameter identification. International Journal of Pure and Applied Mathematics, 52(2):247–265, 2009.

[15] Xueer Bai. Optimization of prognostication model about the spread of Ebola based on SIR model. In 2016 6th International Conference on Machinery, Materials, Environment, Biotechnology and Computer. Atlantis Press, 2016.

[16] Cleo Anastassopoulou, Lucia Russo, Athanasios Tsakris, and Constantinos Siettos. Data-based analysis, modelling and forecasting of the COVID-19 outbreak. PloS ONE, 15(3):e0230405, 2020.

[17] Giulia Giordano, Franco Blanchini, Raffaele Bruno, Patrizio Colaneri, Alessandro Di Filippo, Angela Di Matteo, and Marta Colaneri. Modelling the COVID-19 epidemic and implementation of population-wide interventions in Italy. Nature Medicine, pages 1–6, 2020.

[18] Benjamin Stadlbauer, Andrea Cossettini, Daniel Pasterk, Paolo Scarbolo, Leila Taghizadeh, Clemens Heitzinger, Luca Selmi, et al. Bayesian estimation of physical and geometrical parameters for nanocapacitor array biosensors. Journal of Computational Physics, 397:108874, 2019.

[19] Leila Taghizadeh, Ahmad Karimi, Benjamin Stadlbauer, Wolfgang J Weninger, Eugenijus Kaniusas, and Clemens Heitzinger. Bayesian inversion for electrical-impedance tomography in medical imaging using the nonlinear Poisson–Boltzmann equation. Computer Methods in Applied Mechanics and Engineering, 365:112959, 2020.

[20] Leila Taghizadeh, Ahmad Karimi, Elisabeth Presterl, and Clemens Heitzinger. Bayesian inversion for a biofilm model including quorum sensing. Computers in Biology and Medicine, page 103582, 2019.

[21] Ervin K Lenzi, Luiz Roberto Evangelista, Leila Taghizadeh, Daniel Pasterk, Rafael S Zola, Trifce Sandev, Clemens Heitzinger, and Irina Petreska. Reliability of Poisson–Nernst–Planck anomalous models for impedance spectroscopy. The Journal of Physical Chemistry B, 123(37):7885–7892, 2019.

[22] Ralph C Smith. Uncertainty Quantification: Theory, Implementation, and Applications, volume 12. SIAM, 2013.

[23] Jari Kaipio and Erkki Somersalo. Statistical and Computational Inverse Problems, volume 160. Springer Science & Business Media, 2006.

[24] Walter R Gilks, Sylvia Richardson, and David Spiegelhalter. Markov Chain Monte Carlo in Practice. Chapman & Hall, 1996.

[25] Christian P. Robert and George Cassella. Monte Carlo Statistical Methods. Springer Verlag, 1999.

[26] Malcolm Sambridge and Klaus Mosegaard. Monte Carlo methods in geophysical inverse problems. Reviews of Geophysics, 40(3):1–29, 2002.

[27] Klaus Mosegaard and Malcolm Sambridge. Monte Carlo analysis of inverse problems. Inverse Problems, 18(3):R29, 2002.

[28] Jeffrey S Rosenthal et al. Optimal proposal distributions and adaptive MCMC. Handbook of Markov Chain Monte Carlo, 4, 2011.

[29] Heikki Haario, Marko Laine, Antonietta Mira, and Eero Saksman. DRAM: efficient adaptive MCMC. Statistics and Computing, 16(4):339–354, 2006.

[30] Simon L Cotter, Gareth O Roberts, Andrew M Stuart, and David White. MCMC methods for functions: modifying old algorithms to make them faster. Statistical Science, pages 424–446, 2013.

[31] Heikki Haario, Eero Saksman, and Johanna Tamminen. Adaptive proposal distribution for random walk Metropolis algorithm. Computational Statistics, 14(3):375–396, 1999.

[32] Heikki Haario, Eero Saksman, Johanna Tamminen, et al. An adaptive Metropolis algorithm. Bernoulli, 7(2):223–242, 2001.

[33] Luke Tierney and Antonietta Mira. Some adaptive Monte Carlo methods for Bayesian inference. Statistics in Medicine, 18(1718):2507–2515, 1999.

[34] Peter J Green and Antonietta Mira. Delayed rejection in reversible jump Metropolis–Hastings. Biometrika, 88(4):1035–1053, 2001.

[35] Andrew Gelman, Gareth O Roberts, Walter R Gilks, et al. Efficient Metropolis jumping rules. Bayesian Statistics, 5(599–608):42, 1996.

[36] Henrik Sjödin, Anders F Johansson, Åke Brännström, Zia Farooq, Hedi Katre Kriit, Annelies Wilder-Smith, Christofer Åström, Johan Thunberg, and Joacim Rocklöv. Covid-19 health care demand and mortality in Sweden in response to non-pharmaceutical (NPIs) mitigation and suppression scenarios. MedRxiv https://doi.org/10.1101/2020.03, 20, 2020.

